# Small-quantity lipid-based nutrient supplements for prevention of child malnutrition and promotion of healthy development: Overview of individual participant data meta-analysis and programmatic implications

**DOI:** 10.1101/2021.02.15.21251449

**Authors:** Kathryn G. Dewey, Christine P. Stewart, K. Ryan Wessells, Elizabeth L. Prado, Charles D. Arnold

## Abstract

Small-quantity lipid-based nutrient supplements (SQ-LNS) were designed to provide multiple micronutrients within a food base that also provides energy, protein and essential fatty acids, and were targeted towards the prevention of malnutrition in low- and middle-income countries. Previous meta-analyses have demonstrated beneficial effects of SQ-LNS on child growth, anemia and mortality. To further examine efficacy and effectiveness of SQ-LNS, and explore study-level and individual-level modifiers of the effects, we conducted an individual participant data (IPD) meta-analysis of 14 randomized controlled trials of SQ-LNS provided to infants and young children 6 to 24 mo of age (n > 37,000). We examined growth, development, anemia and micronutrient status outcomes. Children who received SQ-LNS had a 12-14% lower prevalence of stunting, wasting and underweight, were 16-19% less likely to score in the lowest decile for language, social-emotional, and motor development, and had a 16% lower prevalence of anemia and 64% lower prevalence of iron-deficiency anemia, compared to control group children. For most outcomes, beneficial effects of SQ-LNS were evident regardless of study-level characteristics including region, stunting burden, malaria prevalence, sanitation, water quality, duration of supplementation, frequency of contact or average reported compliance with SQ-LNS. For certain outcomes, targeting based on population-level socioeconomic status or undernutrition may be worthwhile, as the benefits of SQ-LNS for iron status, anemia and child development were larger in sub-groups with a greater potential to benefit. A greater impact of SQ-LNS might be possible by co-packaging it with interventions that reduce constraints on response, such as prevention and control of pre-and postnatal infections, improving maternal nutrition, improving health care access, and promotion of early child development. Policy-makers and program planners should consider including SQ-LNS in strategies to reduce child mortality, stunting, wasting, anemia, iron deficiency and developmental impairments. This study was registered at www.crd.york.ac.uk/PROSPERO as CRD42019146592, CRD42020159971 and CRD42020156663.

## Introduction

Millions of infants and young children in low- and middle-income countries are vulnerable to undernutrition and impaired neurobehavioral development (1-3). Globally, 21.3% (144 million) of children under 5 y of age were stunted and 6.9% (47 million) were wasted in 2019 (2). Deficiencies of micronutrients such as iron, zinc, vitamin A and vitamin B_12_ are widespread, particularly among children under 2 y of age, as a consequence of low micronutrient stores at birth, inadequate dietary intake of bioavailable micronutrients, and increased micronutrient requirements due to infection or malabsorption (4, 5). It is estimated that 250 million children under 5 y of age (43%) are at risk of not fulfilling their developmental potential (3), and this is linked to inadequate nutrient intake in early life (6, 7).

The causes of undernutrition and impaired development are complex and multi-factorial (8-13), and thus the impact of interventions that focus solely on improving nutrition may be limited (10, 14). Although dietary interventions by themselves may not be sufficient to eliminate these adverse outcomes, they are a necessary element of strategies aimed at children under 2 y of age, given that inadequate intakes of key nutrients during the complementary feeding period from 6 to 24 mo of age are highly prevalent (8). Improved dietary quality via selection of nutrient-rich complementary foods is the first priority (15, 16), but the cost may be prohibitive for low-income households (17, 18). Various types of fortified products have thus been designed to fill nutrient gaps during the period of complementary feeding, such as fortified blended foods, micronutrient powders (MNP) and lipid-based nutrient supplements (LNS) (19).

During the past 10 y there has been a rapid expansion of research to evaluate the efficacy and effectiveness of LNS in various settings, including meta-analyses of effects of prenatal LNS on birth outcomes (20), and of LNS for children on multiple outcomes (21) including mortality (22). In the meta-analysis of LNS given during the period of complementary feeding by Das et al. (21), 17 trials were included, 13 of which provided small-quantity LNS (SQ-LNS) in at least one arm (the other trials provided larger quantities of LNS). The authors reported positive results for prevention of stunting, wasting and anemia; too few studies were available for child development and micronutrient status outcomes. Although that meta-analysis included some analyses disaggregated by study characteristics, the authors did not conduct analyses stratified by individual-level characteristics.

Since the 2019 meta-analysis by Das et al. (21), additional trials of SQ-LNS have been completed. To examine the current body of evidence relevant to SQ-LNS, and explore reasons for heterogeneity in results, we conducted an individual participant data (IPD) meta-analysis of randomized controlled trials of SQ-LNS provided to infants and young children 6 to 24 mo of age. Our objectives were to 1) generate pooled estimates of the effect of SQ-LNS on outcomes in 3 different domains: a) growth, b) development and c) anemia and micronutrient status, and 2) identify study-level and individual-level modifiers of the effect of SQ-LNS on those outcomes. Identification of subgroups of infants and young children who experience greater benefits from SQ-LNS, or are more likely to respond to the intervention, may be useful in informing the development of public health programs and policies (14). This overview provides a brief history of the development and evaluation of SQ-LNS, a synopsis of the methods of the IPD meta-analyses and the trials included, and a synthesis of the results including discussion of the programmatic and policy implications. Three other papers in this supplement report the detailed methods and results for each of the 3 outcome domains (23-25).

## Development and evaluation of SQ-LNS

In the late 1990s, the first ready-to-use therapeutic food (RUTF) for treating severe malnutrition (Plumpy’nut) was developed, based on embedding micronutrients in a fat-based matrix. This technology allows the product to have a low water activity, which is critical because it inhibits the growth of harmful microorganisms without refrigeration, permitting treatment in outpatient settings. Community-based management of severe malnutrition using RUTF subsequently became widespread. While this revolutionized strategies for *treatment* of malnutrition, there were limited options with regard to effective strategies for *prevention* of malnutrition. To address the latter need, SQ-LNSs were developed based on the same type of food-based matrix used for RUTF (including vegetable oil, peanut paste and milk powder), but using a much smaller quantity of food, typically about 4 teaspoons (∼100-120 kcal) per day (26). The food base provides energy, protein, and essential fatty acids; together with the multiple micronutrients added via fortification, the combination addresses multiple potential nutritional deficiencies.

The target quantity of food in SQ-LNS designed for infants and young children was small for several reasons. First, it is important to avoid displacing breast milk and locally available nutrient-rich foods. The energy needed from complementary foods, assuming average breast milk intakes, is only ∼200 kcal at 6–8 mo, ∼300 at 9–11 mo and ∼550 kcal at 12–23 mo of age. The proportions of these energy needs provided by SQ-LNS are approximately one-half at 6–8 mo, one-third at 9–11 mo, and one-fifth at 12–23 mo, leaving room for other complementary foods in the diet. Second, the small quantity of the daily ration of SQ-LNS makes it likely that the child can consume the entire ration in one day, thereby receiving the intended doses of the micronutrients and essential fatty acids. With a larger quantity, such as medium-quantity LNS (typically 250-500 kcal/d; (26)), a substantial amount may be left unconsumed (27), particularly by infants 6-12 mo of age. Third, the daily ration of SQ-LNS can easily be mixed with other foods (and thus SQ-LNS is considered a type of home fortification product (28)), or consumed as is, allowing for flexibility in feeding practices. Last, the cost of production and transport of LNS, and the feasibility of distribution via platforms such as community health workers, is related to the quantity per recipient, so SQ-LNS is a lower-cost option than other fortified products designed for a larger daily ration.

The first trials evaluating SQ-LNS were published in 2007-2008, and results were promising with regard to prevention of linear growth faltering in Ghana (29) and of severe stunting in Malawi (30). Subsequently, the International Lipid-based Nutrient Supplements (iLiNS) Project (https://ilins.ucdavis.edu/) was funded by the Bill & Melinda Gates Foundation to develop several modified formulations of SQ-LNS (including a version for pregnant and lactating women), conduct large randomized controlled efficacy trials in 3 countries in Africa, conduct socioeconomic studies of SQ-LNS, and coordinate efforts among stakeholders. For the latter objective, the iLiNS Project facilitated the LNS Research Network between 2009 and 2015 to promote exchange of information and experience among researchers and practitioners. These efforts accelerated research on SQ-LNS in a variety of contexts by numerous investigators, both within and outside of the iLiNS Project. This work included studies of acceptability and adherence, trials assessing efficacy and effectiveness for improving a range of different outcomes among infants and young children, as well as pregnant and lactating women, and studies on costs and willingness to pay for SQ-LNS.

Researchers designing trials to evaluate SQ-LNS have recognized that provision of supplements needs to be accompanied by appropriate messages. These include not just information about the use and storage of SQ-LNS but also messages to emphasize recommended infant and young child feeding (IYCF) practices. These messages have typically included promotion of breastfeeding, introduction of complementary foods at 6 mo of age, and recommendations regarding dietary diversity and feeding nutrient-rich complementary foods. These messages have usually been provided to both intervention and control groups, to reinforce the normal IYCF messages already promoted in the study location. Some studies have gone further by providing expanded social and behavior change communication (SBCC) on IYCF. Thus, provision of SQ-LNS generally occurs within an overall context to improve IYCF, not as an isolated intervention solely focused on delivering a product.

## SQ-LNS IPD meta-analysis

### Overview of methods

The IPD meta-analysis presented in the accompanying articles is based on pooled data from 14 randomized controlled trials of SQ-LNS. This work followed best practices for pre-registration, transparency, and reproducibility, with protocols and statistical analysis plans posted online (osf.io/ymsfu and at PROSPERO CRD42019146592, CRD42020159971 and CRD42020156663) (31-34). The trials were identified beginning with those published in a recent Cochrane Review (21), and supplemented with additional studies identified through a systematic review of studies published through September 2019. Investigators who led trials meeting the eligibility criteria shown in **Box 1** were invited to contribute data and participate in the analysis.

#### Box 1. Eligibility criteria for the SQ-LNS individual participant data (IPD) analyses

##### Inclusion criteria

- Randomized controlled trial
- Conducted in low- or middle-income country
- SQ-LNS (< ∼125 kcal/d) provided to the intervention group for at least 3 mo between 6 and 24 mo of age
- At least one trial group did not receive SQ-LNS or other type of child supplementation
- Longitudinal follow-up of each child, or repeated cross-sectional data collection
- Individual children eligible for IPD analysis if:
  - Age at baseline allowed receipt of intervention (supplementation or control group components) for at least 3 mo between 6 and 24 mo of age.
  - For anemia & micronutrient status outcomes, blood samples were collected during the supplementation period or within 3 mo after the study-defined end of supplementation.

##### Exclusion criteria

- LNS was used for treatment, not prevention, of malnutrition (i.e., only children with severe or moderate malnutrition were eligible for the study)
- Conducted in a hospitalized population or among children with a pre-existing disease
- SQ-LNS provision was combined with additional supplemental food or nutrients within a single arm (e.g. SQ-LNS + food rations vs. control), and there was no appropriate comparison group that would allow isolation of the SQ-LNS effect (e.g., food rations alone)

The main comparison of interest was the provision of SQ-LNS (< ∼125 kcal/d, with or without co-interventions) vs. provision of no intervention or an intervention without any type of LNS or other child supplement. For trials with multiple relevant SQ-LNS interventions (e.g., varying dosages or formulations of SQ-LNS in different arms), or that combined provision of SQ-LNS with other non-nutritional interventions (i.e., water, sanitation and hygiene (WASH)), all arms that provided SQ-LNS were combined into one group. All non-LNS arms were combined into a single comparator (“control”) group for each trial [excluding intervention arms that received non-LNS child supplementation, e.g., MNP, fortified-blended food]. For trials that provided both maternal and child LNS, we conducted analyses both with (“all-trials analysis”) and without (“child-LNS-only”) the maternal LNS arms. The main effects did not differ between these two analyses by more than 20% for continuous outcomes or by 0.05 for prevalence ratios, so the results of the all-trials analyses are presented as the principal findings.

We conducted several pre-specified sensitivity analyses: 1) separate comparisons within multi-component intervention trials, such that the SQ-LNS vs. no SQ-LNS comparisons were conducted separately between pairs of arms with the same non-nutrition components (e.g. SQ-LNS+WASH vs. WASH; SQ-LNS vs. Control), 2) exclusion of passive control arms, i.e., control group participants received no intervention and had no contact with project staff between baseline and endline, and 3) exclusion of intervention arms with SQ-LNS formulations that did not include both milk and peanut.

Three types of statistical analyses were conducted to separately investigate 1) full sample treatment effects, 2) effect modification by study-level characteristics, and 3) effect modification by individual-level characteristics. For all 3 sets of analyses, we used a two-stage approach following a complete-case intention-to-treat framework. We used bivariate meta-regression modeling to examine effect modification by study-level characteristics. We modeled effect modification by individual-level characteristics within each study, and then pooled the estimates. Potential effect modifiers examined are shown in **Table 1**.

**Table 1:**
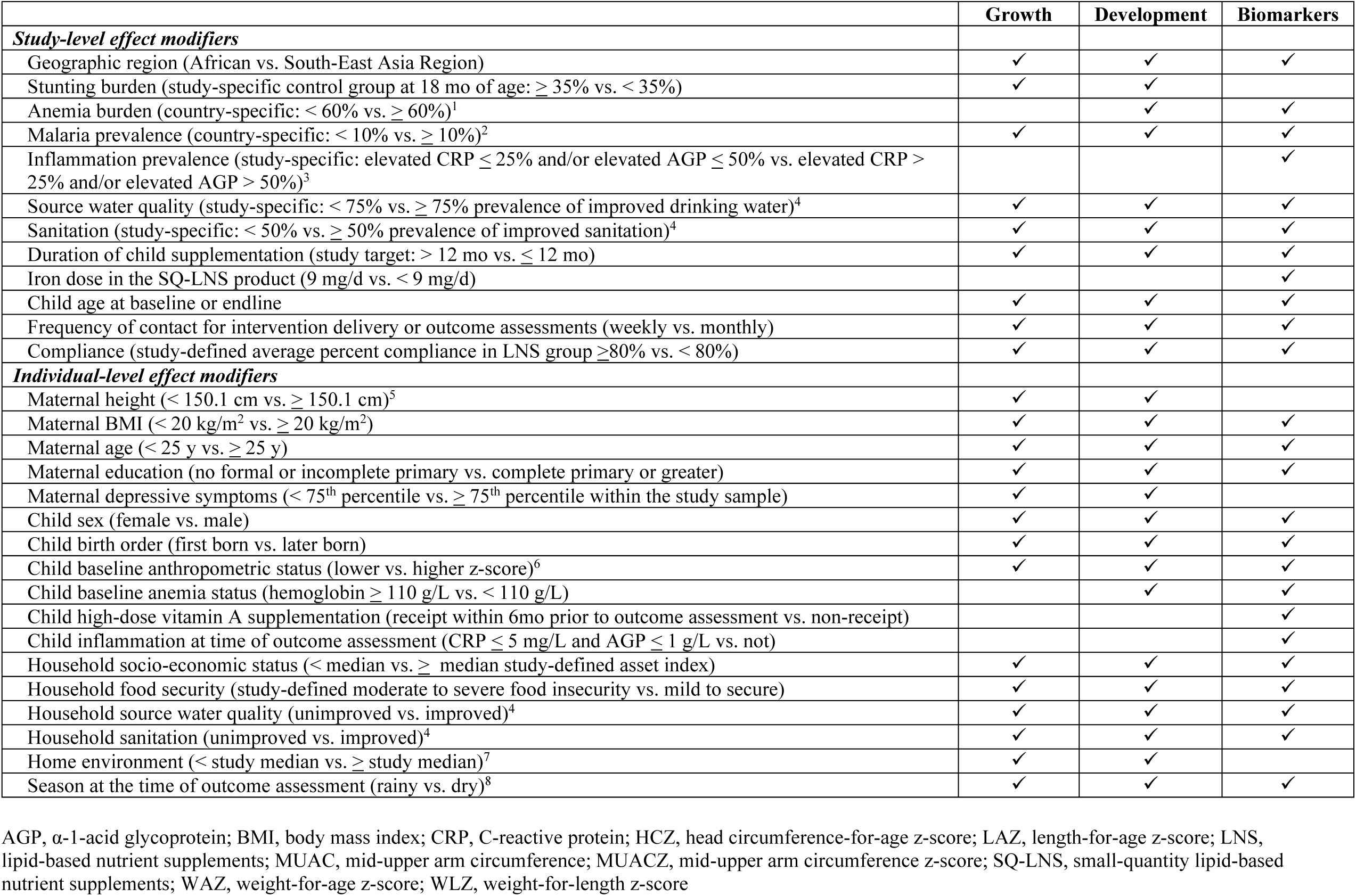
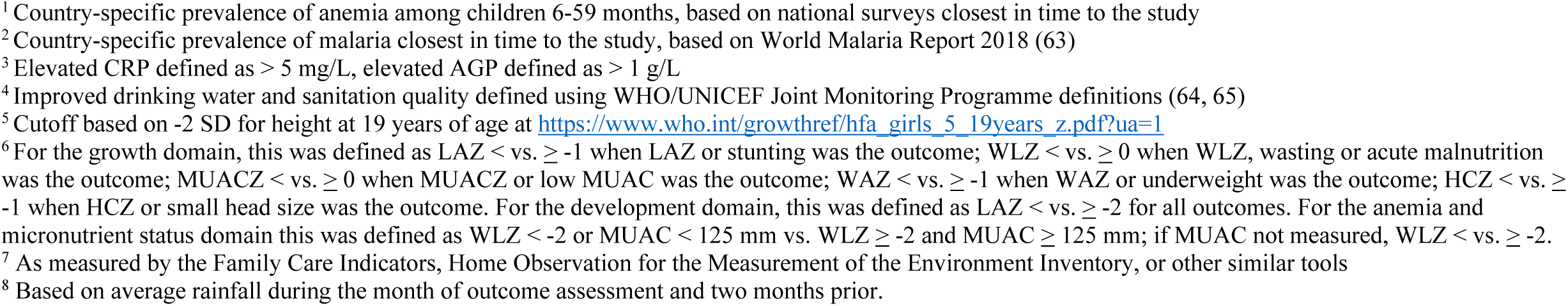
Potential effect modifiers considered within each outcome domain

|  | Growth | Development | Biomarkers |
| --- | --- | --- | --- |
| ***Study-level effect modifiers*** |  |  |  |
| Geographic region (African vs. South-East Asia Region) | ✓ | ✓ | ✓ |
| Stunting burden (study-specific control group at 18 mo of age: $\geq 35\%$ vs. $< 35\%$) | ✓ | ✓ | |
| Anemia burden (country-specific: $< 60\%$ vs. $\geq 60\%$)<sup>1</sup> | | ✓ | ✓ |
| Malaria prevalence (country-specific: $< 10\%$ vs. $\geq 10\%$)<sup>2</sup> | ✓ | ✓ | ✓ |
| Inflammation prevalence (study-specific: elevated CRP $\leq 25\%$ and/or elevated AGP $\leq 50\%$ vs. elevated CRP $> 25\%$ and/or elevated AGP $> 50\%$)<sup>3</sup> | | | ✓ |
| Source water quality (study-specific: $< 75\%$ vs. $\geq 75\%$ prevalence of improved drinking water)<sup>4</sup> | ✓ | ✓ | ✓ |
| Sanitation (study-specific: $< 50\%$ vs. $\geq 50\%$ prevalence of improved sanitation)<sup>4</sup> | ✓ | ✓ | ✓ |
| Duration of child supplementation (study target: $> 12$ mo vs. $\leq 12$ mo) | ✓ | ✓ | ✓ |
| Iron dose in the SQ-LNS product (9 mg/d vs. $< 9$ mg/d) | | | ✓ |
| Child age at baseline or endline | ✓ | ✓ | ✓ |
| Frequency of contact for intervention delivery or outcome assessments (weekly vs. monthly) | ✓ | ✓ | ✓ |
| Compliance (study-defined average percent compliance in LNS group $\geq 80\%$ vs. $< 80\%$) | ✓ | ✓ | ✓ |
| ***Individual-level effect modifiers*** |  |  |  |
| Maternal height ($< 150.1$ cm vs. $\geq 150.1$ cm)<sup>5</sup> | ✓ | ✓ | |
| Maternal BMI ($< 20$ kg/m<sup>2</sup> vs. $\geq 20$ kg/m<sup>2</sup>) | ✓ | ✓ | ✓ |
| Maternal age ($< 25$ y vs. $\geq 25$ y) | ✓ | ✓ | ✓ |
| Maternal education (no formal or incomplete primary vs. complete primary or greater) | ✓ | ✓ | ✓ |
| Maternal depressive symptoms ($< 75^{\text{th}}$ percentile vs. $\geq 75^{\text{th}}$ percentile within the study sample) | ✓ | ✓ | |
| Child sex (female vs. male) | ✓ | ✓ | ✓ |
| Child birth order (first born vs. later born) | ✓ | ✓ | ✓ |
| Child baseline anthropometric status (lower vs. higher z-score)<sup>6</sup> | ✓ | ✓ | ✓ |
| Child baseline anemia status (hemoglobin $\geq 110$ g/L vs. $< 110$ g/L) | | ✓ | ✓ |
| Child high-dose vitamin A supplementation (receipt within 6mo prior to outcome assessment vs. non-receipt) |  |  | ✓ |
| Child inflammation at time of outcome assessment (CRP $\leq 5$ mg/L and AGP $\leq 1$ g/L vs. not) | | | ✓ |
| Household socio-economic status ($<$ median vs. ≥ median study-defined asset index) | ✓ | ✓ | ✓ |
| Household food security (study-defined moderate to severe food insecurity vs. mild to secure) | ✓ | ✓ | ✓ |
| Household source water quality (unimproved vs. improved)<sup>4</sup> | ✓ | ✓ | ✓ |
| Household sanitation (unimproved vs. improved)<sup>4</sup> | ✓ | ✓ | ✓ |
| Home environment ($<$ study median vs. ≥ study median)<sup>7</sup> | ✓ | ✓ | |
| Season at the time of outcome assessment (rainy vs. dry)<sup>8</sup> | ✓ | ✓ | ✓ |

AGP, α-1-acid glycoprotein; BMI, body mass index; CRP, C-reactive protein; HCZ, head circumference-for-age z-score; LAZ, length-for-age z-score; LNS, lipid-based nutrient supplements; MUAC, mid-upper arm circumference; MUACZ, mid-upper arm circumference z-score; SQ-LNS, small-quantity lipid-based nutrient supplements; WAZ, weight-for-age z-score; WLZ, weight-for-length z-score
<sup>1</sup> Country-specific prevalence of anemia among children 6-59 months, based on national surveys closest in time to the study
<sup>2</sup> Country-specific prevalence of malaria closest in time to the study, based on World Malaria Report 2018 (63)
<sup>3</sup> Elevated CRP defined as $> 5$ mg/L, elevated AGP defined as $> 1$ g/L
<sup>4</sup> Improved drinking water and sanitation quality defined using WHO/UNICEF Joint Monitoring Programme definitions (64, 65)
<sup>5</sup> Cutoff based on -2 SD for height at 19 years of age at https://www.who.int/growthref/hfa_girls_5_19years_z.pdf?ua=1
<sup>6</sup> For the growth domain, this was defined as $LAZ < \text{vs. } \geq -1$ when LAZ or stunting was the outcome; $WLZ < \text{vs. } \geq 0$ when WLZ, wasting or acute malnutrition was the outcome; $MUACZ < \text{vs. } \geq 0$ when MUACZ or low MUAC was the outcome; $WAZ < \text{vs. } \geq -1$ when WAZ or underweight was the outcome; $HCZ < \text{vs. } \geq -1$ when HCZ or small head size was the outcome. For the development domain, this was defined as $LAZ < \text{vs. } \geq -2$ for all outcomes. For the anemia and micronutrient status domain this was defined as $WLZ < -2$ or $MUAC < 125$ mm vs. $WLZ \geq -2$ and $MUAC \geq 125$ mm; if MUAC not measured, $WLZ < \text{vs. } \geq -2$ .
<sup>7</sup> As measured by the Family Care Indicators, Home Observation for the Measurement of the Environment Inventory, or other similar tools
<sup>8</sup> Based on average rainfall during the month of outcome assessment and two months prior.

### Trials included

We identified 15 trials that met our inclusion criteria, 14 of which provided individual participant data and were included in the analyses (**Table 2**) (29, 35-48). Investigators for one trial were unable to participate (49). One trial was designed *a priori* to present results separately for HIV-exposed and HIV-unexposed children and is presented as two separate comparisons in all analyses (47, 48). Similarly, the two PROMIS trials in Burkina Faso and Mali each included an independent longitudinal cohort and repeated (at baseline and endline) cross-sectional samples, so the longitudinal and cross-sectional results are presented as separate comparisons for each trial (39, 46).

**Table 2.** Characteristics of the trials included in the SQ-LNS individual participant data (IPD) meta-analyses

| Country | Author | Trial name | Infant SQ-LNS |  | Maternal LNS | Participants |
| --- | --- | --- | --- | --- | --- | --- |
|  |  |  | Age at start (mo) | Duration (mo) |  |  |
| Bangladesh | Christian 2015 (35) | JiVitA-4 | 6 | 12 | N | 4218 |
| Bangladesh | Dewey 2017 (36) | RDNS | 6 | 18 | Y/N | 2478 |
| Bangladesh | Luby 2018 (37) | WASH-B-B | 6 | 18 | N | 4633 |
| Burkina Faso | Hess 2015 (38) | iLiNS-ZINC | 9 | 9 | N | 2626 |
| Burkina Faso | Becquey 2019 (39) | PROMIS-BF | 6 | 12 | N | 2651 |
| Ghana | Adu-Afarwuah 2007 (29) |  | 6 | 6 | N | 194 |
| Ghana | Adu-Afarwuah 2016 (40) | iLiNS-DYAD-G | 6 | 12 | Y | 1040 |
| Haiti | Iannotti 2014 (41) |  | 6-11 | 3-6 | N | 300 |
| Kenya | Null 2018 (42) | WASH-B-K | 6 | 18 | N | 6649 |
| Madagascar | Galasso 2019 (43) | MAHAY | 6-11 | 6-12 | Y/N | 3390 |
| Malawi | Ashorn 2015 (44) | iLiNS-DYAD-M | 6 | 12 | Y | 664 |
| Malawi | Maleta 2015 (45) | iLiNS-DOSE | 6 | 12 | N | 943 |
| Mali | Huybregts 2019 (46) | PROMIS-M | 6 | 18 | N | 2937 |
| Zimbabwe | Humphrey 2019 (47) | SHINE | 6 | 12 | N | 3676 |
| Zimbabwe | Prendergast 2019 (48) | SHINE (HIV-exp) | 6 | 12 | N | 667 |

The 14 trials in these analyses were conducted in Sub-Saharan Africa (10 trials in 7 countries), Bangladesh (3 trials), and Haiti (1 trial), and included a total of 37,066 infants and young children. Most trials began child supplementation with SQ-LNS at 6 mo of age and the intended duration ranged from 6 to 18 mo of supplementation; four trials included intervention arms that also provided SQ-LNS to mothers during pregnancy and the first 6 mo postpartum (36, 40, 43, 44). All trials provided a peanut- and milk-based SQ-LNS in at least one of the arms.

Six trials were conducted within existing community-based or clinic-based programs (36, 39, 41, 43, 46-48); in the other trials, all activities were conducted by research teams. Seven trials provided minimal messaging on IYCF other than reinforcing the normal IYCF messages already promoted in that setting (29, 36, 38, 40, 41, 44, 45), and 7 trials provided expanded SBCC on IYCF that went beyond the usual messaging, either in just the SQ-LNS intervention arms (37, 39, 42, 47, 48) or in all arms including the non-SQ-LNS control arm (35, 43, 46). Three trials included arms with WASH interventions (37, 42, 47, 48). Most trials included an active control arm (i.e., similar contact frequency as for intervention arms) but 3 included only a passive control arm (29, 37, 38).

The 14 study sites were highly diverse in terms of study-level characteristics including stunting burden, malaria prevalence, water quality, sanitation and aspects of study design such as duration of supplementation, frequency of contact and average compliance with SQ-LNS. There was also wide variation within and between studies in maternal, child and household characteristics. This provided heterogeneity for exploration of potential effect modifiers.

### Synthesis of results and programmatic implications

#### Main effects of SQ-LNS

Overall, when combining data from all of the trials, we found significant effects of SQ-LNS across all three outcome domains (**Table 3**). Children who received SQ-LNS had a 12-14% lower prevalence of stunting, wasting and underweight (23), were 16-19% less likely to score in the lowest decile for language, social-emotional, and motor development (24), and had a 16% lower prevalence of anemia and 64% lower prevalence of iron-deficiency anemia (25), compared to control children who did not receive SQ-LNS. These findings add to those of a recently published meta-analysis of many of these same trials reporting a 27% lower risk of mortality between 6 and 24 mo of age (22).

**Table 3.** Relative reductions in adverse outcomes^1^ in meta-analyses of intervention trials providing SQ-LNS to children 6-24 mo of age

| **Growth outcomes** | **Relative reduction (95% CI) %** |
| --- | --- |
| Stunting (LAZ < -2 SD) | 12 (9, 15) % |
| Wasting (WLZ < -2 SD) | 14 (7, 20) % |
| Underweight (WAZ < -2 SD) | 13 (9, 17) % |
| Acute malnutrition (WLZ < -2 SD or MUAC < 125 mm) | 14 (7, 20) % |
| Low MUAC (MUACZ < -2 SD or MUAC < 125 mm) | 18 (11, 25) % |
| Small head circumference (HCZ < -2 SD) | 9 (5, 14) % |
| **Development outcomes<sup>2</sup>** |  |
| Low language development score | 16 (8, 24) % |
| Low motor development score | 16 (8, 24) % |
| Low social-emotional development score | 19 (11, 26) % |
| **Anemia & micronutrient status outcomes** |  |
| Anemia (Hb < 110 g/L) | 16 (13, 19) % |
| Moderate-severe anemia (Hb < 100 g/L) | 28 (24, 32) % |
| Iron deficiency (Ferritin < 12µg/L) | 56 (50, 61) % |
| Iron deficiency anemia (Hb < 110 g/L and ferritin < 12 µg/L) | 64 (56, 70) % |
| Vitamin A deficiency (RBP < 0.70 µmol/L) | 56 (30, 73) % |
| **Mortality** | 27 (11, 41) % |

Hb, hemoglobin; HCZ, head circumference-for-age z-score; LAZ, length-for-age z-score; MUAC, mid-upper arm circumference; MUACZ, mid-upper arm circumference z-score; RBP, retinol binding protein; SQ-LNS, small-quantity lipid-based nutrient supplements; WAZ, weight-for-age z-score; WLZ, weight-for-length z-score
<sup>1</sup> Based on Dewey et al. ((23), growth outcomes), Prado et al. ((24), development outcomes), Wessells et al. ((25), anemia and micronutrient status outcomes), and Stewart et al. ((22), mortality).
<sup>2</sup> Lowest decile defined for each study based on the within-study distribution.

This IPD meta-analysis includes nearly 3 times as many participants as the meta-analysis by Das et al. (21), even though we restricted the analysis to trials that provided SQ-LNS. Our new estimates for wasting and underweight are similar to those of Das et al., but the new estimate for stunting (12% relative reduction) is larger than previously reported (7% reduction). For child development outcomes, Das et al. provided a narrative review of effects, but were not able to generate pooled estimates. With regard to anemia, Das et al. reported a relative reduction of 21%; our 16% reduction is somewhat lower than that, but we defined anemia as hemoglobin < 110 g/L whereas Das et al. examined anemia as defined by trialists. In addition, we report 56-64% reductions in iron deficiency and iron-deficiency anemia, which were not reported in the meta-analysis by Das et al. The larger reduction in prevalence of iron-deficiency anemia as compared to all-cause anemia reflects the complex etiology of anemia; supplementation is likely to prevent only the fraction of anemia attributable to nutritional causes.

The overall effects of SQ-LNS on the growth and development outcomes were modest. However, the effects were generally more consistent, and for some outcomes more substantial, compared to other nutrition interventions for children under 2 y of age, such as nutrition education, micronutrient supplementation or fortification, and fortified blended foods (50-54). Many studies have examined the impacts of behavior change interventions focused on improving complementary feeding practices. A 2018 Cochrane review concluded that such interventions are effective at improving reported feeding practices, but there was insufficient evidence to draw conclusions with respect to effects on growth, development, anemia, or micronutrient status (51). Within our IPD analysis, three trials had direct comparisons of LNS+IYCF behavior change versus IYCF behavior change alone: the JiVitA-4 trial in Bangladesh (35), the PROMIS study in Mali (46), and the Mahay study in Madagascar (43). In 2 of these 3 trials (JiVitA-4 and PROMIS-Mali), the children in the LNS intervention arm had improved growth, hemoglobin and motor development scores compared to children in the IYCF-only arm.

Effects of SQ-LNS on anemia and iron deficiency were similar to those reported in a recent meta-analysis of MNP (18% reduction in anemia and 53% reduction in iron deficiency; (55)). However, that review demonstrated no effects of MNP on child growth and there was insufficient evidence to evaluate effects on child development or mortality.

Compared to interventions promoting responsive care and provision of learning opportunities for young children, effects of SQ-LNS on mean scores for child development were substantially smaller: pooled effect sizes for the former ranged from 0.39 to 0.49 SD higher scores of language, social-emotional, and motor development (56, 57) compared to 0.07 to 0.08 SD higher scores for those domains in the SQ-LNS IPD analysis. However, the former trials did not report estimates for the prevalence of children scoring below a given cut-off in each domain, so we cannot directly compare results with the impact of SQ-LNS on the percentage of children scoring in the lowest decile (relative reductions of 16-19%). Many of the responsive care and learning opportunities studies did not measure indicators of child growth; however, among those that did, there was little evidence of effect (56). Thus, the two types of interventions may be complementary.

#### Effect modification

Each of the accompanying papers presents detailed information on the effect modification results. For most of the outcome domains, effect modification was examined in 3 ways: as the mean difference in the continuous outcome (e.g., LAZ) between SQ-LNS and control groups, as the prevalence ratio between the 2 groups for a binary outcome (e.g., the relative risk of stunting), and as the difference in absolute prevalence between the 2 groups for a binary outcome (e.g., the percentage point difference in prevalence of stunting). We considered all 3 types of outcomes together and in reference to 3 different theoretical scenarios: potential to benefit, potential to respond, and cutoff effects.

##### Potential to benefit

One possibility is that certain subgroups of children may be more likely to benefit from the SQ-LNS intervention, perhaps due to greater nutritional deficits at baseline (14). This is illustrated in **Figure 1A**. The children with greater nutritional or developmental deficits in subgroup a-1 are shown here as being slightly smaller and thinner compared to the children in subgroup a-2. When SQ-LNS is provided, the children in subgroup a-1 may benefit more. Our interpretation in these cases is that there were greater benefits to those who needed it most.

**Figure 1:**
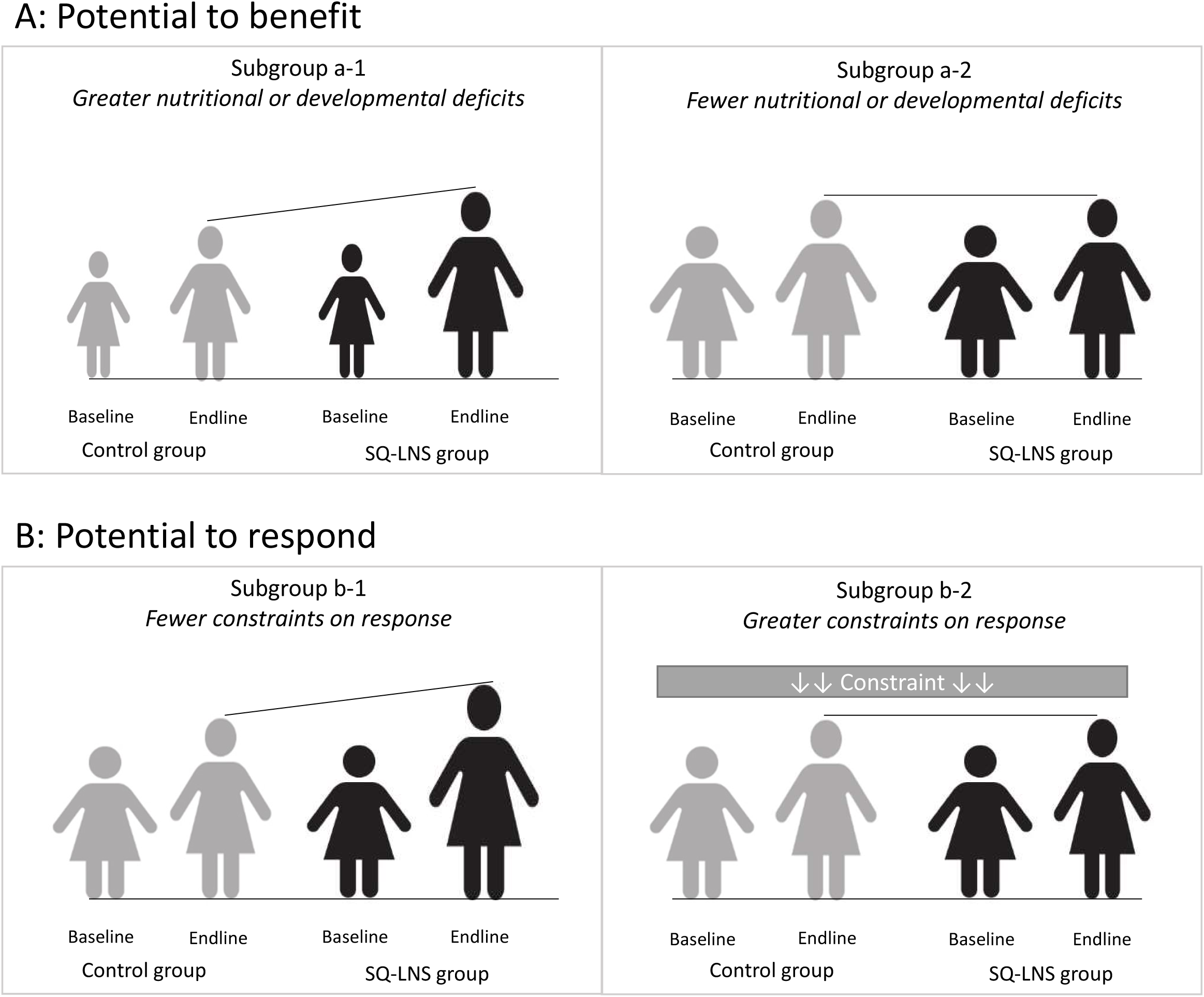
Illustration of the concepts of potential to benefit and potential to respond. A: Subgroup a-1 has greater nutritional deficits, here illustrated as being slightly smaller and thinner than subgroup a-2, and therefore has a greater potential to benefit from a nutritional intervention. B: Subgroup b-1 has a greater potential to respond to a nutritional intervention, due to fewer constraints, as compared to subgroup b-2. SQ-LNS, small-quantity lipid-based nutrient supplements.

##### Potential to respond

Another possibility is that certain subgroups may differ in their potential to respond to the intervention (14), as a result of constraints such as infection or inflammation, caregiver time or resources, or chronic stress. In **Figure 1B**, subgroup b-2 has greater constraints whereas subgroup b-1 has no such constraints. In this case, we might expect to see a greater response to supplementation in subgroup b-1.

##### Cutoff effects

Effect modification results may differ as an artifact of where the continuous outcome distribution falls with respect to the cutoff value for a binary variable. In **Figure 2**, subgroup B has a higher proportion of observations clustered near the cutoff value. Assuming similar shifts in the means of the distributions within both subgroups in response to the intervention, a very different proportion of children cross the cutoff value threshold. In this example, a greater proportion of children in subgroup B would cross that threshold as compared to subgroup A. Thus, there may be statistically significant effect modification for a binary outcome measure, but not for the corresponding continuous outcome measure. In these situations, if there is no significant effect modification for the continuous outcome, we interpret the results as evidence that both subgroups benefit and respond similarly. For this reason, we have not highlighted these examples for further discussion here, but they are described in the accompanying papers (23-25).

**Figure 2:**
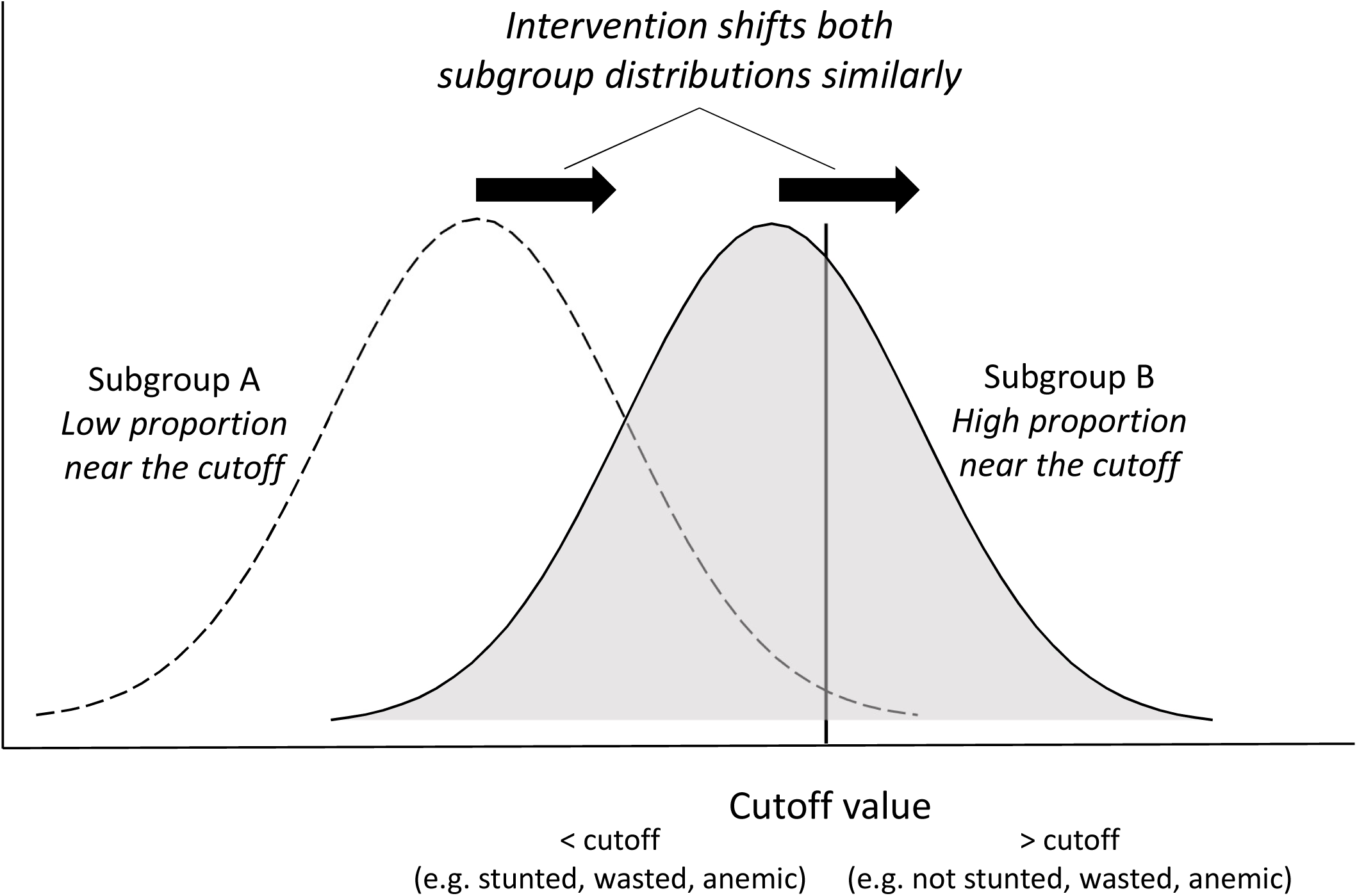
Illustration of the cutoff effect. The intervention shifts both subgroup distributions similarly, resulting in a higher proportion of the individuals in subgroup B crossing the cutoff value threshold. SQ-LNS, small-quantity lipid-based nutrient supplements.

For growth and development outcomes, most of the study-level characteristics and aspects of study design did not significantly modify the effects of SQ-LNS; the exceptions were stunting and anemia burdens, which were related to the impact of SQ-LNS on child development outcomes. For anemia and iron deficiency, several study characteristics (region, anemia burden, malaria prevalence, and prevalence of inflammation) and aspects of study design (duration of supplementation, iron dose in SQ-LNS, and average compliance with SQ-LNS) appeared to modify the effects of SQ-LNS. For all outcomes, there was significant effect modification by individual-level characteristics, but the characteristics that appeared to be important varied considerably depending on the type of outcome. In the paragraphs that follow, we discuss effect modifiers that were common across multiple outcome domains or that might have programmatic implications, using the concepts of potential to benefit and potential to respond described above to interpret the findings.

For two characteristics, child-level acute malnutrition and country-level anemia burden, the results suggested that populations with greater nutritional deficits may have greater potential to benefit from supplementation. Among children acutely malnourished at baseline, plasma ferritin concentration increased by 85% and language, social-emotional, and motor development scores increased by approximately 0.3 standard deviations in the SQ-LNS vs. control groups. In contrast, among children not acutely malnourished, there was a smaller, though still significant impact on plasma ferritin (47% increase) and an increase of only ∼0.1 SD in developmental scores. Similarly, in sites with a higher anemia burden, there were larger differences in mean hemoglobin (+4.9 in SQ-LNS vs. control groups in high anemia burden sites compared to +2.5 g/L in low anemia burden sites), and larger reductions in the prevalence of moderate-to-severe anemia (−13 vs. −4 percentage points) and lowest decile of motor scores (25% vs 12%).

Effect modification by two other characteristics, household socioeconomic status (SES) and study-level stunting burden, suggested that children experiencing greater levels of poverty benefitted more with respect to the effect of SQ-LNS on child development. Such children may be at greater risk of developmental impairment (24) and therefore may have a greater potential to benefit developmentally from supplementation. The mean effects of SQ-LNS on language, social-emotional, and motor development scores in low-SES households were +0.06 to +0.12 SD, compared to little to no effects among children in higher SES households. Similarly, in sites with a high stunting burden, language, social-emotional, and motor development scores were +0.08 to +0.13 SD higher in the SQ-LNS vs. control groups; in contrast, there were no effects on these outcomes in settings with a low stunting burden. While these differences were apparent for the development outcomes, SES and stunting burden generally did not significantly modify the effects of SQ-LNS on growth, anemia, or iron status outcomes.

Child sex was an effect modifier of the effect of SQ-LNS on growth and anemia, with stronger effects among girls than boys. Among girls, SQ-LNS reduced the prevalence of stunting by 16% (vs. 9% among boys), wasting by 21% (vs. 10%), low MUAC by 27% (vs. 7%), small head size by 15% (vs. 4%) and anemia by 18% (vs. 13%), though the latter difference was likely due to the cutoff effect. Girls had higher mean anthropometric z-scores and hemoglobin concentrations than boys, which suggests that they did not have a greater potential to benefit from the supplementation. Rather, they may have had a greater potential to respond. Boys are at greater risk of morbidity and mortality in early life and may be more vulnerable to environmental stressors (58), which could constrain their response to a nutrition intervention. However, there were significant positive effects of SQ-LNS on mean z-scores for growth, hemoglobin concentration and indicators of iron status among both boys and girls. Additionally, child sex did not modify the effect of SQ-LNS on any indicators of child development. Thus, both boys and girls benefitted from the intervention.

Child birth order and maternal age both modified the effects of SQ-LNS on more than one outcome domain. Effects of SQ-LNS were greater among later-born children (i.e., those with at least one older sibling) than among first-born children with regard to stunting (13% vs 9% relative reduction among later-born vs. first-born children, respectively), underweight (17% vs 6% reduction), low MUAC (23% vs 5% reduction), anemia (12 vs 7 percentage point reduction), and several continuous outcomes (mean WAZ, MUACZ, motor and fine motor scores, hemoglobin, and ferritin concentrations). Similarly, effects of SQ-LNS were greater among children born to older mothers than among those born to younger mothers with regard to anemia (17% vs 13% relative reduction) and mean motor and fine motor scores. Birth order and maternal age are positively correlated, so it is difficult to disentangle which factor is the most likely driver of these differences. Later-born children have at least one older sibling with whom they may compete for resources, making them potentially more vulnerable to malnutrition. In fact, we observed lower mean hemoglobin concentration and a greater prevalence of stunting and underweight among later-born (vs first-born) children in the control groups in the IPD analysis, suggesting that they have a greater potential to benefit from nutritional supplementation.

Maternal education and depressive symptoms also appeared to modify the effect of SQ-LNS on growth and development, but in seemingly disparate ways. Among children whose mothers had higher education or less depressive symptoms, there were greater effects of SQ-LNS on growth. This may reflect a greater potential to respond to the intervention. Such mothers may have had greater autonomy and agency, and therefore may have been better able to adhere to advice regarding the recommended frequency or dosage of supplementation. By contrast, among children whose mothers had lower education, there were larger beneficial effects of SQ-LNS on child development. Other studies have reported that maternal education is associated with developmental delays (59), and therefore these children may have lagged behind their peers and had greater room for improvement, with greater potential to benefit from an intervention.

Effect modification by season of assessment was significant for wasting and iron status, although the direction of the relationship differed by outcome. Specifically, there were greater effects of SQ-LNS on the prevalence of wasting among children assessed during the dry season (22% reduction) compared to children assessed during the rainy season (8% reduction), which may reflect a greater potential to respond in the dry season due to a lower burden of infections. On the other hand, effects of SQ-LNS on iron status were greater when it was measured during the rainy season, with an increase of 74% in ferritin concentration and a decrease of 63% in the prevalence of iron deficiency relative to the control groups. In the dry season, a benefit of SQ-LNS was still apparent (44% increase in ferritin and 47% reduction in the prevalence of iron deficiency), but the effect size was smaller. These results do not suggest that SQ-LNS interventions should be targeted to one season or another, but they have important implications with regard to interpreting results from evaluations, particularly if such studies have not been conducted longitudinally across a full calendar year. It is also important to note that the data included in the IPD meta-analysis were generally based on cross-sectional outcome assessments, which do not capture the multiple episodes of wasting/acute malnutrition, micronutrient deficiency, or anemia that may occur throughout the study period. In the PROMIS study in Mali, for example, the longitudinal prevalence of acute malnutrition was much higher than the cross-sectional prevalence, and the SQ-LNS intervention reduced the former by 29% but had no significant impact on the latter (46).

Finally, greater effects of SQ-LNS on iron status and anemia were observed among children without inflammation at the time of assessment. This may reflect a greater potential to respond to a nutrition intervention among children without inflammation, who are likely to experience less inhibition of iron absorption or sequestration of circulating iron (60, 61). Despite this difference, significant positive effects of SQ-LNS on these outcomes were apparent in both sub-groups, with and without inflammation.

## Conclusions

In summary, the evidence suggests that there are important benefits of SQ-LNS for child survival, growth, anemia, iron status, and child development. It must be noted that SQ-LNS is not a stand-alone intervention, as it should always be accompanied by messaging to reinforce IYCF recommendations, including a diverse diet with healthy foods from the key food groups. Nonetheless, it appears to play a protective role when access to nutrient-rich foods is limited for economic or other reasons. Of course, the benefits of SQ-LNS must be weighed against the potential benefits of alternative interventions and the costs of delivering each type of intervention. At present, however, we are not aware of any other intervention that has a demonstrated impact on all of the outcomes mentioned above.

The IPD meta-analysis included >37,000 children from a wide range of settings. For most outcomes, beneficial effects of SQ-LNS were evident regardless of region, stunting burden, malaria prevalence, sanitation, water quality, duration of supplementation, frequency of contact or average reported compliance with SQ-LNS. Moreover, 6 of the 14 trials were conducted within existing community-based or clinic-based programs, so the evidence represents the continuum from efficacy to effectiveness trials. The sensitivity analyses demonstrated very similar results, indicating that the findings are robust. One limitation was that Bangladesh was the only country represented in the Southeast Asia Region and Haiti was the only country represented in Latin America and the Caribbean, so additional data from countries outside of Africa would be valuable.

The effect modification results suggest that for certain outcomes, targeting on the basis of population-level socioeconomic status or burden of undernutrition may be worth considering, as the benefits of SQ-LNS for iron status, anemia and child development were larger in sub-groups who had a greater potential to benefit from the intervention. The results also suggest that a greater impact of SQ-LNS might be obtained by co-packaging it with interventions that alleviate constraints on response, such as the prevention and control of pre-and postnatal infections; improving maternal nutrition; improving access to health care, including mental health care for women; and promoting early child development interventions that promote responsive caregiving.

The selection of intervention(s) for a given population should be based on the needs of the population, the goals of the program, as well as cost:benefit considerations. Although the latter is outside of the scope of this and the accompanying papers, work is under way to understand these economic dimensions. In the meantime, we recommend that policy-makers and program planners consider including SQ-LNS in the mix of interventions to reduce child mortality, stunting, wasting, anemia, iron deficiency and developmental impairments. It is one of the few interventions that can help to achieve multiple Sustainable Development Goal targets simultaneously, as well as support the three pillars of the United Nations’ 2016-2030 Global Strategy for Women’s, Children’s and Adolescents’ health (62): surviving (ending preventable deaths), thriving (ensuring health and well-being) and transforming (expanding enabling environments).

## Data Availability

Data described in the manuscript, code book, and analytic code will not be made available because they are compiled from 14 different trials, and access is under the control of the investigators of each of those trials.

## Acknowledgments

We thank all of the co-investigators, collaborators, study teams, participants and local communities involved in the trials included in these analyses. These trials benefitted from the contributions of many partner organizations, including: icddr,b (JiVitA-4, Rang-Din Nutrition Study and WASH Benefits trial in Bangladesh); the World Food Program (JiVitA-4 trial in Bangladesh); the Health District of Dandé and the relevant local health-care authorities (iLiNS-ZINC trial in Burkina Faso); AfricSanté and Helen Keller International (PROMIS trials in Burkina Faso and Mali); Ministry of Public Health and Population (Haiti trial); Innovations for Poverty Action and the Kenya Medical Research Institute (WASH-Benefits trial in Kenya); Unité Programme National de Nutrition Communautaire, Government of Madagascar, and World Bank Health and Nutrition and Population Global Practice (MAHAY trial in Madagascar); the Ministry of Health and Child Care in Harare, Chirumanzu and Shurugwi districts, and Midlands Province (SHINE trial in Zimbabwe); the International Lipid-based Nutrient Supplements Project Steering Committee (iLiNS Project trials); and Nutriset (for development of SQ-LNS). We thank Emily Smith for advice on IPD analysis methods.

The authors’ responsibilities were as follows—KGD and CPS: drafted the manuscript with input from other coauthors; All authors read, contributed to, and approved the final manuscript.

Supported by Bill & Melinda Gates Foundation grant OPP49817 (to KGD). All authors report no conflicts of interest.

## Abbreviations

AGP: α-1-acid glycoprotein
BMI: body-mass index
CRP: C-reactive protein
Hb: hemoglobin
HCZ: head circumference z-score
iLiNS: International Lipid-based Nutrient Supplements Project
IPD: individual participant data
IYCF: infant and young child feeding
LAZ: length-for-age z-score
LNS: lipid-based nutrient supplement
MNP: multiple micronutrient powder
MUAC: mid-upper arm circumference
MUACZ: mid-upper arm circumference z-score
RBP: retinol binding protein
RDNS: Rang-Din Nutrition Study
RUTF: ready-to-use therapeutic food
SBCC: social and behavior change communication
SES: socio-economic status
SQ-LNS: small-quantity lipid-based nutrient supplements
WASH: water sanitation and hygiene
WAZ: weight-for-age z-score
WLZ: weight-for-length z-score

